# The impact of reactive mass vaccination campaigns on measles outbreaks in the Katanga region, Democratic Republic of Congo

**DOI:** 10.1101/19003434

**Authors:** Sebastian Funk, Saki Takahashi, Joel Hellewell, Kartini Gadroen, Isidro Carrion-Martin, Marit van Lenthe, Katiana Rivette, Sebastian Dietrich, W. John Edmunds, M. Ruby Siddiqui, V. Bhargavi Rao

## Abstract

The Katanga region in the Democratic Republic of Congo (DRC) has been struck by repeated epidemics of measles. In many of the affected health zones, reactive mass vaccination campaigns were conducted in response to the outbreaks. Here, we attempted to determine how effective campaigns were in curtailing a large outbreak in 2015. Using a model of measles transmission we compared observed case numbers to a counterfactual of no campaigns, by first fitting a model to the data including the campaigns and then re-running this without vaccination. Focusing on eight of the 68 health zones in the Katanga region, we estimated the reactive campaigns to have reduced the size of the outbreaks by approximately 21,000 (IQR: 16,000–27,000; 95% CI: 8300–38,000), or 21% (IQR: 17%–25%, 95% CI: 9.3%–34%) of possible measles cases. There was considerable heterogeneity in the impact of campaigns, with campaigns startgin earlier after the start of an outbreak being more impactful. We further sought to establish whether the spatial pattern of the outbreak could have been determined in advance to help prioritise areas for vaccination campaigns and speed up the response. The best predictors of outbreak size among all the health zones were vaccination coverage derived from cluster surveys and outbreak size in 2010-13. This, combined with the fact that the vast majority of reported cases were in under-5 year olds, would suggest that there are systematic issues of undervaccination. If this was to continue, outbreaks would be expected to continue to occur in the affected health zones at regular intervals, mostly concentrated in under-5 year olds. Taken together, our findings suggest that while a strong routine vaccination regime remains the most effective means of measles control, it might be possible to improve the effectiveness of reactive campaigns by considering predictive factors to trigger a more targeted vaccination response.

## Introduction

There have been repeated outbreaks of measles in the Democratic Republic of Congo (DRC). The Katanga region (formerly known as Katanga province) is in the southeast of the country bordering Zambia and comprises the provinces of Haut-Katanga, Haut-Lomami, Lualaba and Tanganyika. It has experienced large periodic measles outbreaks, such as in 2006–07, 2010–13 [1, 2]. In response to these, reactive mass vaccination campaigns have been conducted to protect those assumed to be at risk both within the outbreak area and beyond.

Standard measles epidemic responses include reinforcing measles surveillance in affected areas, providing free care to reduce measles mortality, and reactive vaccination campaigns in order to control measles transmission. In collaboration with the World Health Organization (WHO) Regional Office for Africa (AFRO) and the United Nations Children’s Fund (UNICEF), Médecins Sans Frontières (MSF) supported the Ministry of Health to respond to various measles outbreaks including two major measles outbreaks in the Katanga region. Firstly, in 2010–13, a measles epidemic was reported with over 96,000 suspected cases reported, 77% of which occurred in children under 5 years of age, and more than 1400 deaths [2]. In 2011, in response to the ongoing epidemic, MSF vaccinated more than 1.8 million children 26 of the 68 health zones in the Katanga region [1]. Secondly, in February 2015, a new measles epidemic started in Katanga, DRC, lasting the whole year and resulting in over 40,000 cases and more than 400 deaths in 2015 [3]. MSF responded with the standard epidemic responses including a reactive vaccination campaign in order to stop measles transmission during epidemics, targeting more than 25 health zones.

The time interval between the outbreak starting in different parts of Katanga and the vaccination response implemented varied. Previously, modelling studies in Niger have demonstrated that even late vaccination intervention in response to an outbreak could prevent a large number of cases, though early intervention will always have a larger impact [4, 5, 6, 7]. However, this may be context-specific and vary with local epidemiology and outbreak patterns. The response to the Katanga outbreak provides an opportunity to retrospectively study the effectiveness of the campaigns conducted in mitigating excess morbidity. More generally, important lessons could be learned about the relationship between response times and effectiveness of campaigns, and how campaign targets could be selected in the future to ensure greatest impact.

We studied the 2015 measles outbreak and responsive mass vaccination campaigns conducted as part of the standard epidemic response to assess whether the most-affected areas could have been predicted from information on previous out-breaks and administrative or otherwise estimated vaccination coverage. We further investigated the outbreak in several health zones using a mathematical model of measles transmission, to quantify the impact of vaccination campaigns that were conducted in those areas.

## Methods

### Data sources and cleaning

Suspected measles cases (WHO definition) from 2010–16 were collated from the integrated disease surveillance (IDS) system, described previously in [2]. These data are split into age groups 1-4 years and 5 years and over, at the level of health zones. The database did not contain any information on cases under the age of 1 year.

Administrative coverage data from 2009-16 collected by the Ministry of Health was available as the number of doses administered per week was collected at the level of health zones, separated into age groups 9-11 months and 12-23 months.

Population denominators were extracted from the coverage data. Since the last census in DRC prior to this study had been done in 1981, these numbers are subject to considerable uncertainty.

We further used vaccination coverage estimates from a previous study [8]. These used data collected as part of the Demographic and Health Survey (DHS) in 2013– 14, extrapolated from geo-located information on children’s vaccination status from vaccine cards and parental recall. We averaged the estimates by month of age to arrive at the proportion of under-5 year olds that were unvaccinated, that is had received no dose of measles-containing vaccine.

Information on reactive mass vaccination campaigns conducted in 2015 was extracted from MSF reports. The total number of vaccine doses administered was collated at the level of health zones, and at various temporal resolutions from days to a single number of doses delivered for a whole campaign.

### Factors that could predict outbreak size

We tested the predictability of outbreaks from demographic factors and outbreak and vaccination history in a negative binomial Generalized Linear Model with logarithmic link. Robust standard errors and p-values were calculated using the *sandwich R* package [9, 10]. The number of suspected cases reported during the 2015 outbreak at the health zone level was modelled as a function of health zone population size, the number of cases in the 2010–13 outbreak, MoH administrative and estimated vaccination coverage.

### Modelling measles with mass vaccination campaigns

We modelled measles transmission at the level of health zones using a stochastic transmission model with a fixed time step of 2 weeks, corresponding to the generation time of measles [11]. At each time step *t*, the number of new infections in health zone *i, I*_*it*_ was drawn from a negative binomial distribution with mean *λ*_*it*_*S*_*i*(*t−*1)_ and shape *m*, allowing for overdispersion of transmission, or superspreading [12]:

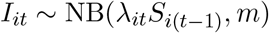

where *S*_*i*(*t−*1)_ and *I*_*i*(*t−*1)_ are the number of people susceptible and infected, respectively, at time *t−*1, and *λ*_*it*_ is the force of infection experienced by susceptibles in health zone *i* at time *t*:

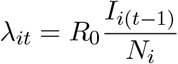

where *N*_*i*_ is the population size of health zone *i, R*_0_ is the basic reproduction number.

When a mass vaccination campaign was conducted, the number of susceptible people immunised was calculated by multiplying the number of doses administered with the proportion of the population still susceptible *S*_*it*_*/N*_*i*_, and a campaign efficiency factor *e*_*i*_, estimated as part of the inference procedure described below. This factor comprises both vaccine efficacy and the efficiency in targeting susceptible children, which were not identifiable separately. With a perfect vaccine and random distribution, this would take a value of 1. If vaccines were preferentially given to susceptibles, it would take values of greater than 1 (subject to vaccine efficacy). If vaccines were preferentially given to already immune children, it would take values of less than 1.

During a two-week span, half of vaccinations were modelled to be administered before transmission occurred and half afterwards. While the measles vaccine takes 2 weeks to come into effect, it provides potentially high level of protection from 72 hours after administration [13, 14, 15]. We therefore assumed that vaccination starts to fully immunise a child instantaneously.

For the counterfactual scenarios of how the outbreaks would have evolved without a reactive mass vaccination, we simulated the model from the time of the mass vaccination campaigns, but without reducing the number of susceptibles as a consequence of vaccination. We then drew samples from the joint distribution of trajectories and observations, to obtain alternative trajectories of observed cases. To evaluate the impact of the campaigns, we calculated the reduction in the number of cases observed in each of the trajectories. If this yielded a negative difference (i.e., if random sampling yielded alternative trajectories with more cases than the observed ones), we treated the impact as 0 (i.e., same number of cases in both scenarios).

### Selection of health zones for fitting and estimating populations

The health zones selected for the dynamic model were ones that reported more than 10 cases in at least one week in 2015 and had a reactive mass vaccination campaign with the number of doses delivered and results from a follow-up coverage survey available. A total of eight health zones were modelled, including the one that saw most cases (Malemba-Nkulu, 8856 reported cases) and 7 of the 13 health zones with most cases in 2015: Ankoro (3910), Kinkondja (2773), Mukanga (2723), Bukama (2632), Songa (928) and Kabalo (904).

Since a large proportion of cases was found in children (77% in 1-to-5 year olds, with no further age-breakdown available), and none of the vaccination campaigns targeted over-15 year olds, we modelled measles transmission to be occurring exclusively in under-5 year olds. The relevant population sizes were estimated as the number of doses administered in the vaccination campaigns divided by the coverage estimated from concurrent vaccination surveys. Where vaccination campaigns were limited to under-5 or under-10 year olds, we estimated the total population size under 15 as 2.72 or 1.39 times the estimated population size, respectively, based on multipliers used for estimating the sizes of age groups in the administrative coverage data provided.

### Model fitting and counterfactual scenarios

The model was fitted simultaneously to the eight selected health zones. The likelihood of observing bi-weekly incidence *D*_*it*_ in health zone *i* at time *t* was taken to follow a negative binomial distribution with fixed overdispersion *ϕ*.

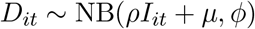

where *ρ* is the proportion of cases that is reported, $*µ* is the rate of background reporting of measles, either due to cases that were not part of the epidemic or misclassification, for example of rubella cases, and *ϕ* is the reporting overdispersion.

The value of the basic reproduction number *R*_0_, the efficacy of mass vaccination *e*_*i*_, mean reporting rate *ρ*, background reporting rate *m*, observation overdispersions, the proportion immune *r*_*i*0_ in health zone *I* and the mean number of individuals infectious *I*_*i*0_ at the first data point with at least 10 cases in health zone *i* (taken to be the start of the time series), were all estimated as part of the inference procedure, as well as likely trajectories of the state variables. The reporting rate *ρ*_*i*_ and initial number infectious *I*_*i*0_ was allowed to vary between health zones. The prior distribution on the mean reporting rate was weakly informed by a coverage survey that was conducted in Kabalo. The initial proportion immune *r*_*i*0_ was estimated with a mean and lower bound given by the vaccination coverage per health zone *v*_*i*_ estimated in [8]. Informed or regularising prior distributions of the parameters to be estimated are shown in Table 1.

**Table 1:**
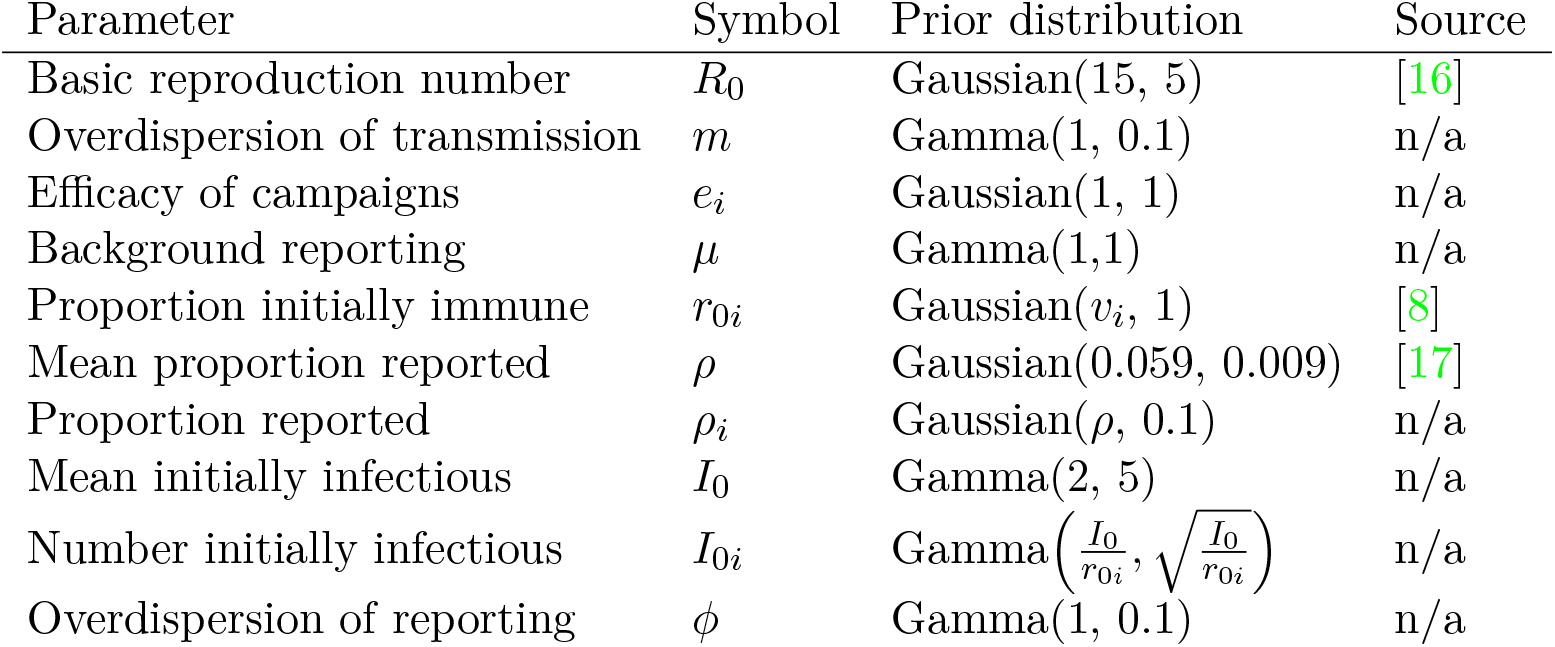
Prior distributions of parameters used in the transmission model. The distribution of the basic reproduction number was truncated at a lower bound of 0. The propotion initially immune was truncated to be between v_i_ and 1. The mean and actual proportions reported were truncated to be between 0 and 1. The number initially infectious were trunctedat a lower bound of 0.

The model was fitted to the data using a particle filter in combination with Metropolis-Hastings Markov chain Monte Carlo (pMCMC) with the *libbi* software library [18] as implemented in the *RBi* package using the statistical software *R* [19, 20]. The number of particles and proposal distribution was adapted using the *RBi*.*helpers* package [21], before the pMCMC sampler was run to generate 4096 samples after thinning, with 262,144 particles. The inference pipeline was run on an Nvidia Tesla P100 16GB NVLink GPU.

## Results

### Outbreak size

In total, 40,562 cases and 485 deaths were reported in the Katanga region over the course of the year (case-fatality ratio: 1.2%). The majority of cases were reported from Haut-Lomami (23,984, 59%) and Tanganyika (12,110, 30%) provinces, with the outbreak in Tanganyika peaking significantly later than the one in Haut-Lomami (Fig. 2). Of the 68 health zones, 16 reported over 90% of cases (Fig. 1).

**Figure 1:**
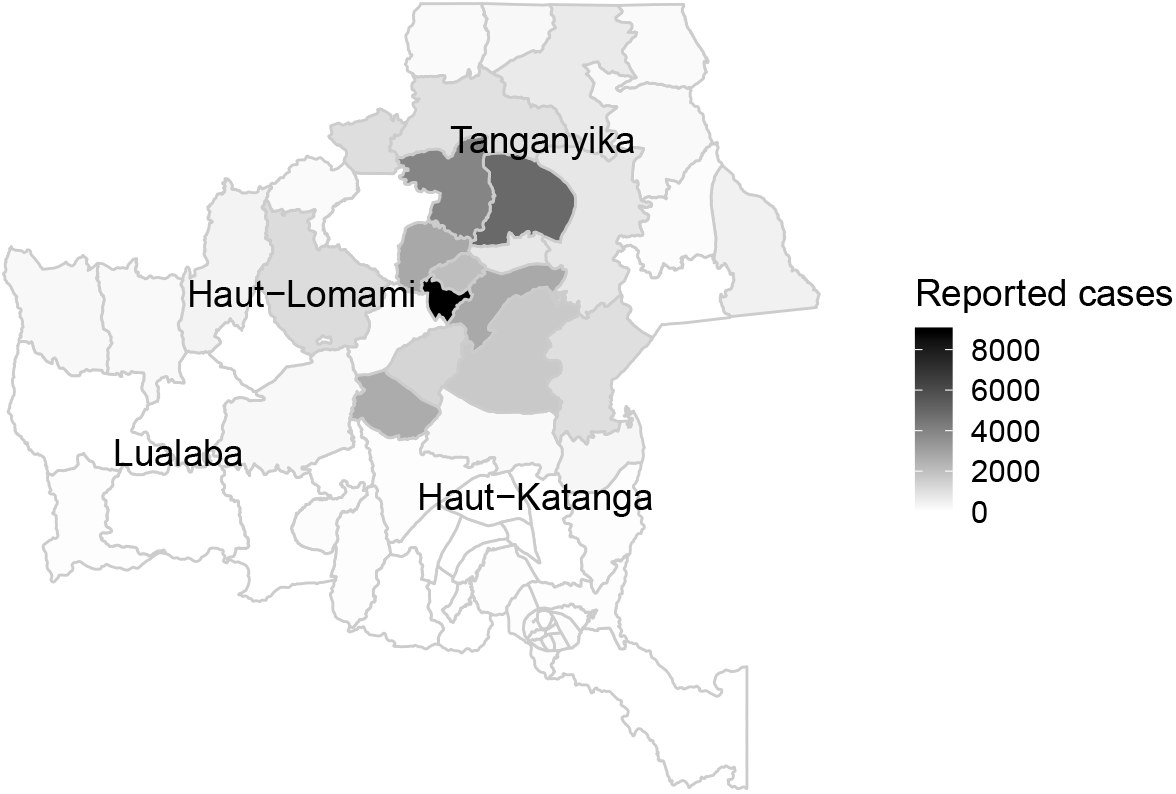
Number of cases by health zone in the Katanga region, 2015.

**Figure 2:**
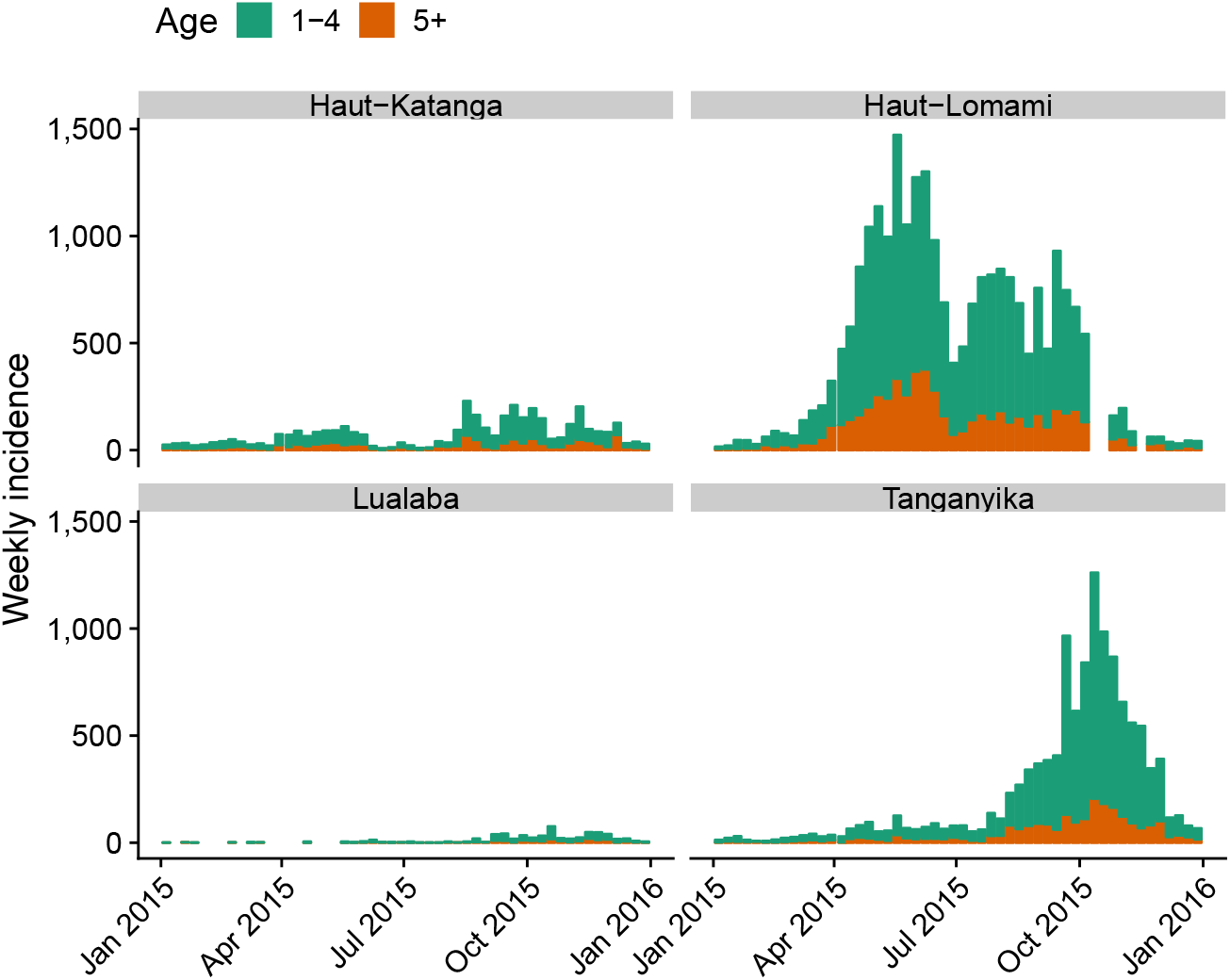
Number of cases by age group and province in Katanga, 2015.

### Predictability of outbreak size

There was a positive correlation between reported incidence in the 2010–13 outbreak and the 2015 outbreak (Pearson’s r=0.31, p=0.01, Fig. 3). All the health zones with more than 10 cases per 1000 in 2015 (Malemba-Nkulu, Kinkondja, Manono, Ankoro, Lwamba, Mitwaba, Mukanga, Bukama) had also reported more than 5 cases per 1000 in 2010–13.

**Figure 3:**
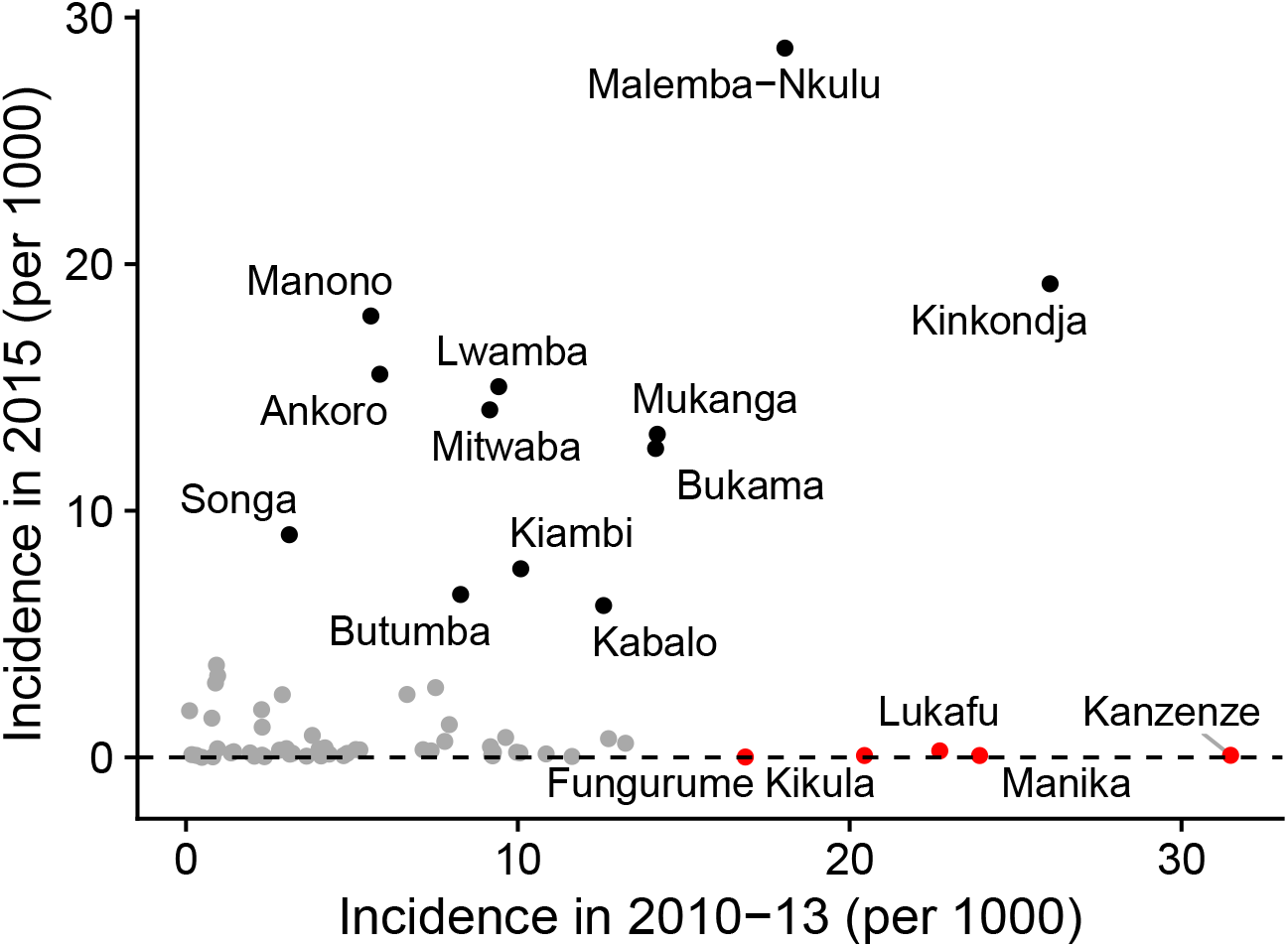
Incidence (number of cases divided by estimated population size) in 2010– 13 vs 2015. Health zones with more than 5 cases per 1000 in 2015 are indicated in black, and other health zones with more than 10 cases per 1000 in 2010–13 in red.

Further, there was a positive correlation of reported incidence in 2015 and admin-istrative vaccination coverage, and a negative correlation with coverage as estimated from DHS data (Fig. 4).

**Figure 4:**
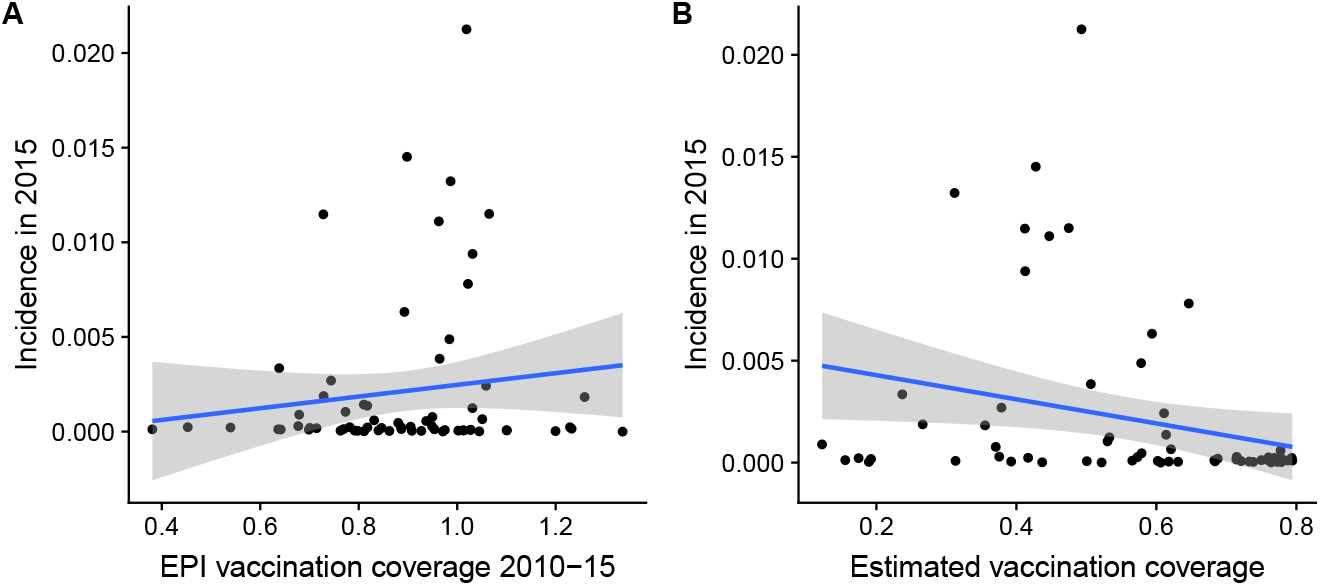
Vaccination coverage versus reported incidence (number of cases divided by estimated population size) in 2015. Linear trends are indicated by blue lines, with 95% confidence intervals indicated in grey. A) Mean vaccination coverage in 2010–15 as measured by the EPI programme. B) Vaccination coverage estimated from DHS data.

Combining these factors and population size in a regression model confirms these correlations, with coefficients corresponding to the number of cases in 2010–13 and vaccination coverage estimated by DHS as strongest predictors of the number of cases that occurred in 2015 (Table 2). Population size and routine vaccination coverage as measured by the EPI programme did not have a strong influence on the number of cases in 2015. Correlation between model predictions and true number of cases was 0.3 (95% CI 0.1-0.5, p=0.01, Fig. 5).

**Table 2:** Regression coefficients for model of case numbers in 2015, with lower and upper 95% confidence interval limits.

| Coefficient | Estimate | p-value | Lower limit | Upper limit |
| --- | --- | --- | --- | --- |
| (Intercept) | 5.7 | <0.001 | 5.4 | 6.1 |
| Population size | 0.1 | 0.8 | -0.4 | 0.6 |
| Number of cases 2010–13 | 0.8 | <0.001 | 0.2 | 1.3 |
| Mean EPI coverage 2010–15 | 0.3 | 0.09 | -0.1 | 0.7 |
| DHS coverage estimate | -1.3 | <0.001 | -1.8 | -0.9 |

**Figure 5.**
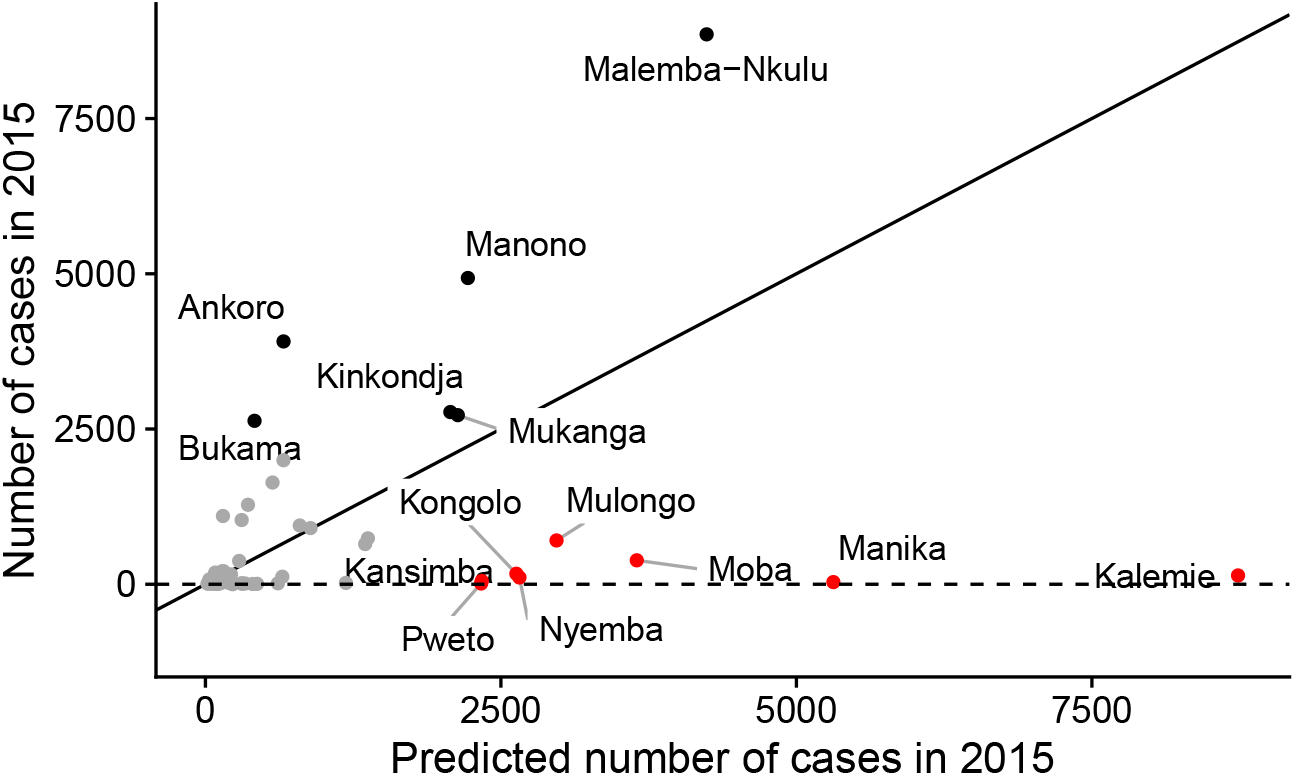
Predictions from the regression model vs. true number of cases. As in Fig. 3, health zones with more than 5 cases per 1000 in 2015 are indicated in black, and other health zones with more than 10 cases per 1000 in 2010–13 in red.

**Table 3:** Summary of posterior estimates.

| Parameter | Symbol | Posterior mean | (IQR) |
| --- | --- | --- | --- |
| Basic reproduction number | $R_0$ | 4.3 | (4.0–4.5) |
| Overdispersion of transmission | $m$ | 0.17 | (0.14–0.2) |
| Efficacy of campaigns (mean) | $e_i$ | 0.34 | (0.14–0.48) |
| Background reporting | $\mu$ | 1.4 | (1.0–1.7) |
| Proportion initially immune (mean) | $r_{0i}$ | 0.55 | (0.49–0.62) |
| Number initially infectious (mean) | $I_{0i}$ | 66 | (46–78) |
| Proportion of cases reported (mean) | $\rho_i$ | 0.24 | (0.19–0.29) |
| Overdispersion of reporting | $\phi$ | 0.044 | (0.022–0.061) |

To further investigate the relationships underlying the results, we tested an ad-Coefficient Estimate p-value Lower limit Upper limit ditional model variant, where we distinguished the four provinces comprising the Katanga region in the model, to determine whether effects were being identified at the fine level of the health zone or the coarser province level. In that case, province as a categorical explanatory variable in the regression replaced some of the predictive value both of the number of cases in 2010–13 (regression coefficient 0.4, p=0.05) and the coverage estimate from DHS data (-1.1, p*<*0.001), but both retained predictive value, the coverage estimate strongly so. This suggests that some predictive value of case numbers in 2010–13, and strong predictive value of the coverage estimate was retained at the lower level of the health zone.

### The impact of mass vaccination campaigns

To investigate the impact of the mass vaccination campaign in more detail, we fitted a dynamic model to the case trajectories in 8 health zones (Fig. 6). We estimated a basic reproduction number of 4.3 (mean; interquartile range, IQR: 4.0– 4.5) and an average reporting rate of 24% (IQR: 19%-29%), corresponding to a total of 77,000 (IQR: 73,000–81,000; 95% CI: 66,000–91,000) estimated cases from 19,079 reported cases in the 8 health zones. On average, 55% (IQR: 49%-62%) of under-5 year olds were estimated to have been immune before the outbreak. The estimated campaign efficacy factor ranged from 0.21 (IQR: 0.09–0.31) in Kinkondja to 0.59 (IQR: 0.33–0.83) in Ankoro.

**Figure 6:**
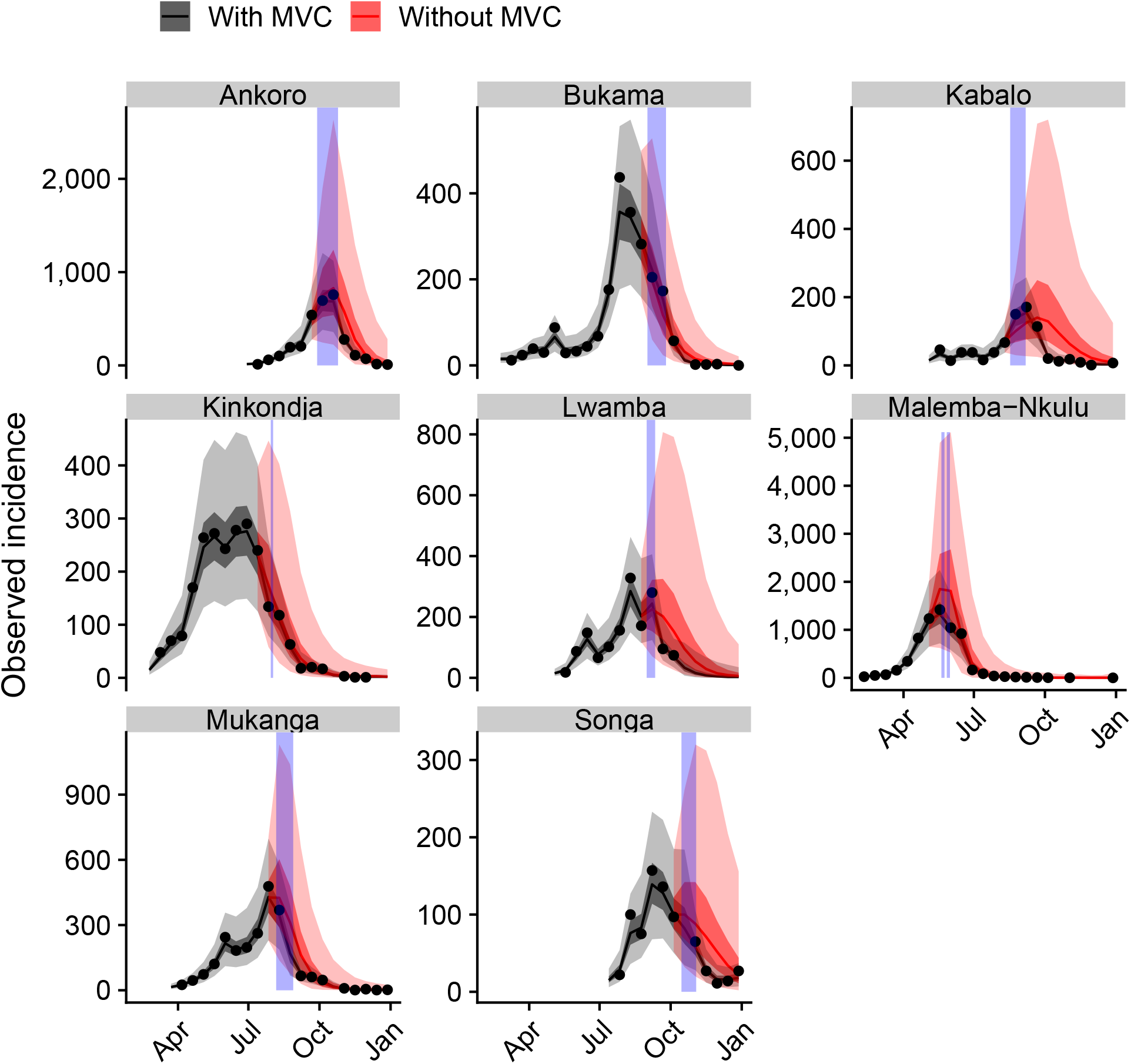
Model fits (black) to the 2015 data and counterfactual scenarios without mass vaccination campaigns (red). The data are shown as black dots, and periods of mass vaccination campaigns as blue vertical bars. Median fitted trajectories are shown as lines, 50% (dark grey) and 95% (light grey) credible intervals as shades.

In total, we estimate that 21,000 (IQR: 16,000–27,000; 95% CI: 8300–38,000) cases were averted by the vaccination campaigns in the seven health zones analysed, corresponding to relative reduction in case load of 21% (IQR: 17%–25%, 95% CI: 9.3%–34%). Of the approximately 250,000 doses delivered to under-5 year olds in the 8 health zones, we estimated 22,000 (IQR: 17,000–26,000, 95% CI: 11,000– 37,000) or 9.2% (IQR: 7.2%–11%, 95% CI: 4.5%–15%) of administered doses went to susceptible children.

There was heterogeneity in impact between health zones. The greatest absolute impact achieved by a mass vaccination campaign in the health zones investigated was in Malemba-Nkulu with 6800 (IQR: 4000–9100; 95% CI: 0–17,000) cases averted with 26,208 doses, while the greatest relative impact was in Kabalo with a 33% (IQR: 17%–54%; 95% CI: 0%–73%) reduction in case load from an estimated 20,727 doses (Table 4). On the other hand, only 230 (IQR: 0–810; 95% CI: 0– 2400) or 2.4% (IQR: 0%–11%; 95% CI: 0%–29%) of cases were estimated to have been averted in Bukama from an estimate 31,400 doses. Speed of implementation of the mass vaccination campaign (or shorter delay to implementation) was highly correlated with a greater relative reduction of cases (Pearson’s p = -0.85, p=0.008).

**Table 4:** Absolute and relative impact of mass vaccination campaigns in different health zones. Estimates shown are posterior means. The delay shown in the last column is the number of weeks between the start of the outbreak (end of the first two-week period with more than 10 cases) and the beginning of the vaccination campaign.

| Health zone | Doses<br>(est.) | Cases<br>averted | (IQR, 95% CI) | Relative<br>reduction | (IQR, 95% CI) | Delay<br>(weeks) |
| --- | --- | --- | --- | --- | --- | --- |
| Ankoro | 26,199 | 4800 | (2200–7300, 0–12,000) | 24% | (13%–37%, 0%–55%) | 11 |
| Bukama | 34,100 | 230 | (0–810, 0–2400) | 2.4% | (0%–11%, 0%–29%) | 25 |
| Kabalo | 20,727 | 3000 | (1000–4700, 0–9100) | 33% | (17%–54%, 0%–73%) | 13 |
| Kinkondja | 20,792 | 510 | (0–970, 0–2800) | 5.5% | (0%–12%, 0%–29%) | 20 |
| Lwamba | 44,148 | 3400 | (870–5400, 0–12,000) | 21% | (6.7%–35%, 0%–61%) | 14 |
| Malembe-Nkulu | 46,330 | 6800 | (4000–9100, 0–17,000) | 23% | (16%–31%, 0%–47%) | 14 |
| Mukanga | 30,133 | 2200 | (670–3500, 0–6800) | 15% | (5.7%–25%, 0%–44%) | 17 |
| Songa | 19,660 | 970 | (240–1500, 0–3300) | 19% | (6.2%–32%, 0%–54%) | 11 |

## Discussion

In spite of repeated strategic and reactive vaccination campaigns, large measles outbreaks continue to occur in Katanga, DRC, causing significant morbidity and mortality. Strategies to mitigate the burden of measles are urgently needed. Here we conducted both predictive and retrospective modelling of the measles outbreaks in Katanga in 2015, with the aim to evaluate the impact of the vaccination response as well as potential for improvement.

The predictability of outbreaks is related to the quality of the available data. We found little relationship between reported administrative vaccination coverage and observed incidence. In fact, there was a small positive correlation, that is more cases occur where vaccination uptake as indicated by the EPI programme is higher. This could be because high routine vaccination rates might be an indicator of surveillance quality and therefore case reporting. At the same time, Strategic Immunisation Activities were conducted across Katanga after the 2011 outbreak [22]. We did not have access to any details of these campaigns, which may have been targeted at areas with low reported vaccination rates, thus raising immunity in those health zones. Not all of the suspected cases included in this study may have been measles and instead have been misdiagnoses due to rubella or other causes of rash [23]. While we included a parameter for misclassification in the modelling analysis, this is difficult to identify and may be an underestimate. Lastly, there is uncertainty around the population estimates used as denominator when estimating coverage, as high rates of migration and urban growth make existing data quickly outdated.

Vaccination rates as estimated from cluster surveys as part of the DHS programme, on the other hand, were well correlated with case data, with higher vaccination rates corresponding to lower case burden. These estimates encompass all vaccination activities and not just routine immunisation, and they do not suffer from denominator issues caused by uncertainty in the population sizes within health zones.

Reconstructing the outbreak with a mathematical model of the case trajectories suggested that reactive mass vaccination campaigns reduced the case load substantially, and more so the earlier it was implemented. We estimated that tens of thousands of susceptibles were immunised during those campaigns and, consequently, tens of thousands of cases averted in under-5 year olds. While the estimated over-all proportion of doses that went to susceptibles may appear low at approximately 10%, this must be seen in the context of conducting vaccination campaigns during ongoing outbreaks, where part of the population may already have been infected and thus naturally immunised. In all health zones, we estimated that vaccines were preferentially given to immune children, who may have been immunised through routine vaccination, been targeted in previous campaigns, or infected and acquired natural immunity during the ongoing or previous outbreaks. At the same time, the estimated 21,000 cases averted correspond to a reduction in burden of over 20%. In the health zones modelled, the case-fatality ratio in the reported data was 1.2%, suggesting that around a hundred infant lives were probably saved by the campaigns.

Our transmission model suffered from several limitations. We did not have access to an age breakdown of cases older than 5 years, and information on under-1 year olds was missing completely. Because of this, we only modelled transmission in under-5 year olds. At 77% of reported cases, it seems safe to assume that transmission in under-5 year olds was driving the outbreaks. The estimated basic reproduction number of 4.3 (IQR: 4.0–4.5) is small in comparison with other settings, possibly because transmission does not occur in school-like settings with close mixing of large numbers of children, but rather households and communities affecting children before they reach school age.

The estimated impact of the campaigns might have been greater if cases averted in over 5-year olds had been taken into account. We further ignored any spatial progression of the outbreak or connectivity between health zones and modelled each area in isolation. In reality, mass vaccination campaigns that reduced cases in one area may well have prevented subsequent cases in nearby areas in other health zones. Lastly, we assumed constant reporting rates. If, on the other hand, reporting quality changes between regions or over time, it would affect our fits which would interpret these changes as changes in transmission rather than reporting.

In spite of enormous efforts, measles is proving difficult to control in Katanga. On the 10th June 2019, the DRC Ministry of Health officially declared a new measles outbreak in 23 out of the 26 provinces of DRC, with initial cases for this outbreak reported in late 2018. This new measles outbreak coincided with an ongoing Ebola outbreak in the North Kivu and Ituri provinces of DRC which had begun in August 2018. There have been suggestions that the diversion of resources and attention towards the Ebola response may have reduced the healthcare capacity required to respond to a surge in measles cases [24]. Although at the time of writing, the health zones most affected by the measles outbreak were outside the area where Ebola was mostly concentrated, it has been shown during the 2013–16 outbreak in West Africa that reduced vaccination services as a result of an Ebola outbreak can have a severe impact on measles circulation [25, 26, 27].

The ability to partly predict the case load in 2015 from outbreaks in 2010–13 at the province level suggests that there might be underlying problems in the provision of routine immunisation services that did not change in the intervening time. At the end of outbreaks as big as the ones occurring in Katanga, not many children are left susceptible, whether a mass vaccination campaign has been conducted or not. The fact that another big outbreak could happen so soon after the last suggests a rapid increase in susceptibles that have not been served by the routine vaccination programme, and strengthening this should be a priority. At the same time, it is clear that the mass vaccination campaigns only prevent part of the observed cases, partly because of unavoidable delays in confirming an outbreak and launching a campaign. Preventive strategies based on predictive models have a potential to have a much greater impact if they can prevent outbreaks altogether, but their use is based on the predictive potential of the models used. We found that vaccination estimates based on a spatial model applied previously to vaccination survey data was a good predictor of outbreak size at the relatively fine level of health zones. There is enormous promise in using such estimates to guide strategic immunisation activities and close any existing gaps in immunity. As has been proven many times over, it is only through strong and comprehensive routine vaccination, supplemented by strategic campaigns where necessary, that sustained measles control and, ultimately, elimination can be achieved.

## Ethics

This research fulfilled the exemption criteria set by the MSF Ethics Review Board (ERB) for a posteriori analyses of routinely collected clinical data and thus did not require MSF ERB review.

## Data Availability

All data are available with an R package that can be used to reproduce the results. This is available at https://github.com/sbfnk/measles.katanga

## Acknowledgements

The authors would like to thank colleagues from the Democratic Republic of Congo, including Chefs de la Division Provinciale de la Santé from the following regions: Patrick M’piongo (Haut Lomami), Gerard Mwabu Mbumbu (Lualaba), Jerry Kyungu Kibanza (Tanganyika) and Jean-Marie Kafwimbi (Haut Katanga). In addition we thank colleagues from MSF who shared their operational data and who facilitated sharing of historical data, especially Narcisse Mukembe and Florent Uzzeni (MSF Switzerland), Liliana Palacios (MSF Spain) and Axelle De La Motte (MSF France). Finally we like to acknowledge and thank Marion Dols (MSF Netherlands) for her earlier work on measles within MSF operations.

This work was supported by the Wellcome Trust [grant number 210758/Z/18/Z] and the UK Economic and Social Research Council [grant number ES/P010873/1].

## Data statement

All code and data used to produce the results are available in a public repository at https://github.com/sbfnk/measles.katanga.

## Author statement

All authors attest they meet the ICMJE criteria for authorship.

